# Estimating the potential impact of the UK salt reduction targets on cardiovascular health outcomes in adults: a modelling study

**DOI:** 10.1101/2025.04.16.25325974

**Authors:** Lauren Bandy, Ben Amies-Cull, Madison Luick, Linda J. Cobiac, Susan A. Jebb, Peter Scarborough

## Abstract

**Background:** Excessive sodium intake is responsible for three million deaths a year globally. The UK is one of 64 countries to have a salt reduction programme to help reduce the population’s sodium intake. It is a voluntary scheme with 108 category-specific salt content targets for the grocery and out of home sectors. The aim of this study was to estimate the potential impact of these targets on cardiovascular outcomes and healthcare costs for UK adults.

**Methods:** Long-term health modelling was based on the adult population in England. Changes in salt intake (grams/day) were estimated using consumption data from the National Diet and Nutrition Survey 2017/18. Impact on ischaemic heart disease (IHD) and stroke, QALYs and healthcare costs were estimated using PRIMEtime, a proportional multistate lifetable model.

**Results:** If the salt reduction targets set for 2024 were met, then salt intake would reduce from 6.06g/day (95% CI 5.18 – 6.31) to 4.94g/day (4.73 – 5.15), a reduction of 1.12g/day (1.05 – 1.20).

This would lead to 103,000 (UI 41,000-161,000) fewer cases of IHD and 25,000 (10,000 – 39,000) fewer cases of stroke over 20 years. A modelled 243,000 (94,000-383,000) QALYs would be saved with a net saving of £1.00 billion (£0.35-1.73 billion)) to the NHS over the remaining lifetime of the adult population.

**Discussion:** Reformulation of products to meet the targets could result in substantial reductions in cardiovascular disease without changes in dietary behaviours. Policymakers should consider options to strengthen salt reduction policies, including effective systems for monitoring and enforcement.

## Introduction

Excess dietary salt is responsible for three million deaths a year globally (1). There is strong evidence that reducing dietary sodium leads to dose-dependent declines in both systolic and diastolic blood pressure (2–4) which are key risk factors for cardiovascular diseases (5,6). WHO recommends that salt intake should not exceed 5g/day and that salt reduction is one of the most cost-effective measures a country can take to improve population health (7). The UK government’s salt reduction programme first began in 2003 and consisted of a set of voluntary targets as well as product labelling, a public and political awareness campaign and monitoring (8). Since then, the campaigning element of the salt reduction programme has diminished, but the Government has published a series of updated targets for industry in 2009, 2011, 2014, 2017 and mostly recently in 2020 to be met by 2024 (9). Sales-weighted mean salt content and maximum salt content targets were set for 84 grocery food categories, such as bread, cheeses, meats and snacks, and, for the first time, 24 food categories consumed out of the home, including chips, burgers, curries and pizza (9). The aim of these targets has been to encourage manufacturers to gradually reduce the salt content of everyday foods so that consumers do not notice a change in taste, therefore reducing the impact on consumer preference or product sales.

Previous studies have reported that initially these targets were successful at reducing sodium consumption. Population salt intakes in England fell by 15%, from 9.5g/day in 2003 to 8.1g/day in 2011 (10). Over the same time period, a fall in the population’s blood pressure of 3.0/1.4 mmHg (diastolic/ systolic) was observed, contributing to an estimated 42% and 40% reduction in stroke and IHD mortality respectively (10).

More recently, however, there have been concerns that progress at reducing dietary sodium intake has stalled (11). Dietary survey data shows that the UK population’s salt intake has begun to increase, from 8.1g/day in 2011 to 8.4g/day in 2017-18 (12). Evidence suggests that between 2015 and 2020, there has been little change in the salt content of grocery foods, including bread, cheese and ready meals (13). Between 2015 and 2020, the mean salt content of grocery foods in the UK fell by just 0.05g/100g, with little to no change in leading grocery categories such as bread and ready meals (13). A 2020 report on the food industry’s progress towards the 2017 targets by Public Health England (PHE) showed mixed results, with only half of the grocery targets being met (14). With a new government and salt targets due for review, it is pertinent to examine the potential benefits of salt reduction for population health.

The aim of this study was to estimate the potential impact the 2020 UK salt reduction targets would have on average UK salt intakes if they had been met in full by the food industry by 2024, and to estimate their potential impacts on cardiovascular outcomes and healthcare costs at the population level.

## Methods

We modelled a scenario estimating the potential changes in population salt intake, ischaemic heart disease, stroke and healthcare costs that could be achieved by the government’s 2020-2024 salt reduction target programme for foods consumed both in and out of the home in the UK. Ethics committee approval was not required.

### Estimating changes in salt consumption

The National Diet and Nutrition Survey (NDNS) is a rolling annual cross-sectional survey that collects data on food consumption and nutrient intakes of a representative sample of approximately 1000 individuals (around 500 children and 500 adults) using repeated food diaries for a 3-4 day period to account for day-to-day variation (15). Data are published by the UK Government and were accessed through the UK Data Service (16). Individual-level microdata on the volume of food products consumed (g/day) and their corresponding salt content data (g/100g) were taken from NDNS Year 11 (2018-19). Each food product was matched to one of the 108 categories covered in the salt reduction programme, based on its category name and product description. The salt target category names and the number of foods from NDNS matched to each category is given in Supplementary Table 1. NDNS survey weighting values were applied to increase the representativeness of the sample to the UK population. The baseline daily salt intakes (g/day) for adults by sex were calculated by summing salt intake across all food categories for pre-prepared and packaged foods, including those purchased from takeaways (i.e. Foods that fell outside the salt reduction target categories, or were described as ‘homemade’, were not included in baseline salt intakes in this paper.).

**Table 1:** Daily salt intake for adults aged with 95% confidence intervals.

|  | Female Population | Male Population | Population Average |
| --- | --- | --- | --- |
| Baseline salt intake, g/day<br>Mean (95% CI) | 5.26 (4.99 – 5.54) | 6.98 (6.59 – 7.36) | 6.06 (5.18 – 6.31) |
| Intervention salt intake, g/day<br>Mean (95% CI) | 4.33 (4.08 – 4.58) | 5.63 (5.32 – 5.95) | 4.94 (4.73 – 5.15) |
| Salt reduction, g/day<br>Mean (95% CI) | 0.93 (0.85 – 1.02) | 1.34 (1.22 – 1.47) | 1.12 (1.05 – 1.20) |
| Salt reduction as proportion of<br>baseline salt intake<br>Mean (%) (95% CI) | 16.76 (15.78 –<br>17.75) | 18.33 (17.17 –<br>19.49) | 17.49 (16.73 –<br>18.25) |

The salt content (g/100g) of each food was then re-populated with the category-specific target values given in the salt reduction programme. Foods that were described as ‘takeaway’ were categorised according to the out of home sector targets as given in the salt reduction programme. The targets for the out of home sector are given per serving size, not per 100g, therefore it was assumed the reported ‘total grams consumed’ variable in NDNS was equivalent to serving size. The scenario daily salt intake (g/day) for each adult was calculated by summing salt intake across all recorded food categories using these re-populated target salt values for foods.

We corrected for underreporting in the dietary recall questionnaire of the NDNS by assuming the pattern of underreporting in the dietary recall questionnaire was equal across all foods. We did not adjust for the salt target categories that could not be matched in the NDNS data. A correction factor was calculated as the ratio between the published mean estimated salt intake from the NDNS England Sodium Survey 2018/19 (12) and baseline mean salt intakes estimated in this study, by age group (19-34, 35-49 and 50-64 years) and sex as published in the National Sodium Survey. The Sodium Survey is based on urinary sodium, so the ratio represents the proportional difference between what is reported in the dietary survey (whether from food products or added salt) and what can be inferred they are consuming (whether from food products or added salt). The estimates of salt consumption at baseline and scenario were multiplied by these factors and differences calculated between the two to capture the change to salt consumption related to the targets, assuming that people are under-reporting consumption of target and non-target food categories equally.

### Estimating impacts on disease burden and healthcare costs

The impact of the scenario changes to mean salt intakes on the disease burden of cardiovascular disease and ischaemic stroke was modelled using the PRIMEtime model. This is a proportional multistate lifetable model that has been described fully elsewhere (17,18). A brief description of how it operates follows. Firstly, the risk module quantifies the impact of the change in risk factor on the incidence of diseases. This is done by calculating the Population Impact Fraction for each disease outcome, which represents the proportional difference in disease incidence related to a change in a related risk factor. This is done by estimating the impact of a change to salt consumption on systolic BP using the conversion factor of 5.80mmHg (millimetres of mercury) (2.45 – 9.15mmHg) per 100 millimoles of sodium (equating to 2.30g of sodium or 5.88g of salt) (4) and calculating confidence intervals for scenario changes to BP using 10,000 iterations of Monte Carlo analysis. Second, the disease module separately quantifies the impact of the change in disease incidence to disease burden over time, and finally, the lifetable module compiles these disease burden impacts into a total population burden. The population is structured by age and sex, and is simulated to age across the chosen time horizon simultaneously with baseline and scenario disease incidence, allowing the difference between the two to be calculated. The 2020 population of the UK was used from the Office for National Statistics, baseline disease epidemiology from the Global Burden of Disease 2019 with secondary modelling using DISMOD-II and blood pressure-disease relationships were quantified using Relative Risks from a prospective cohort study (19).

The reduction in salt intake was implemented linearly over the 4 years and the baseline comparator was for no change to mean salt intakes. Disease utility values were taken from a UK study (20) and unit healthcare costs calculated as previously described for PRIMEtime (17,18). A discount rate of 1.5% was used for disease burden and healthcare costs, in line with the DHSC and NICE guidance for public health interventions (21). A time lag of 5 years was set between changing BP and the increase to disease incidence rates (18).

A flow chart summarising the methods is given below (**Figure 1**).

**Figure 1:**
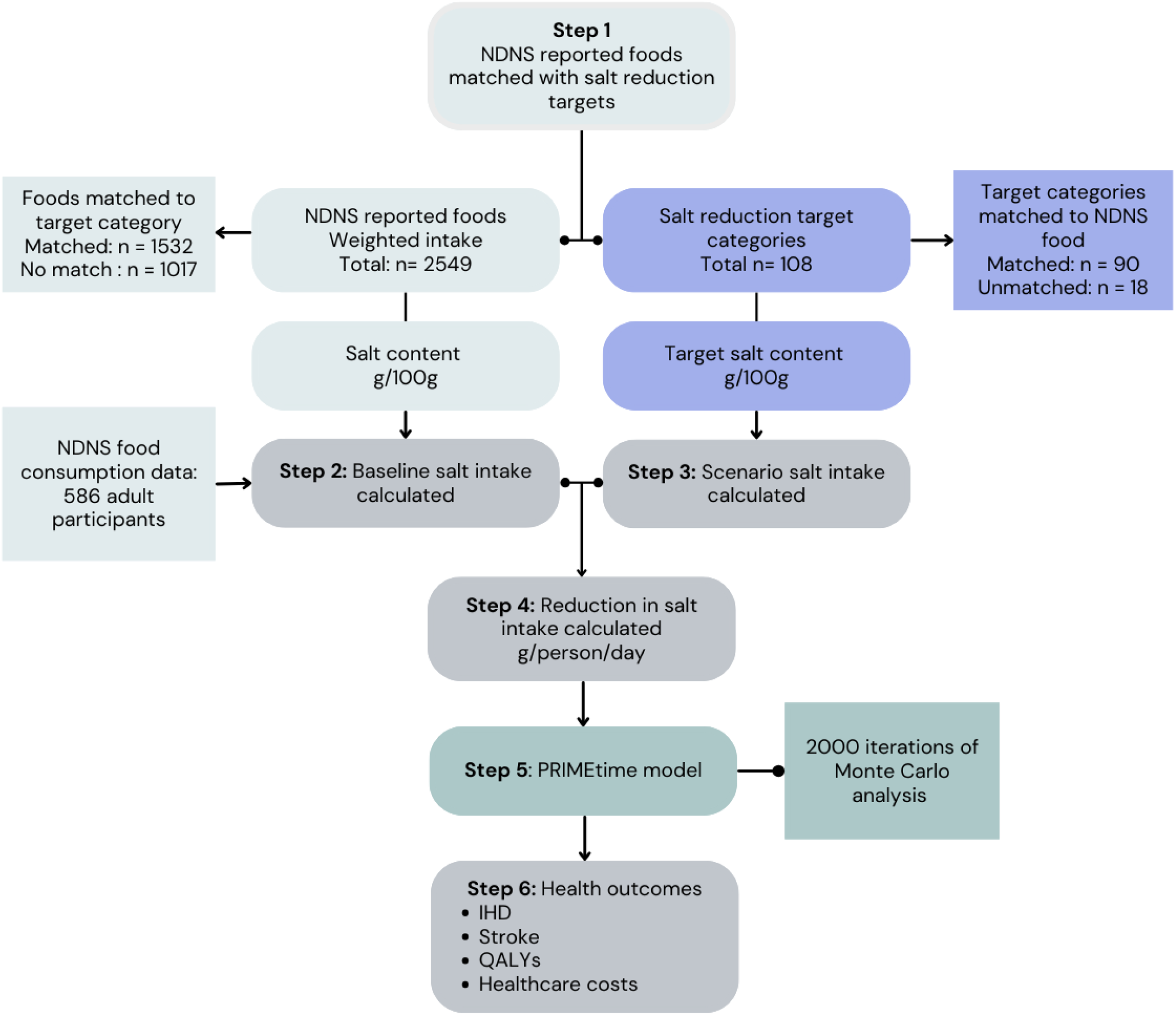
Summary of methods

### Key assumptions

The scenario was modelled assuming that the programme had no impacts on consumer behaviour. Specifically, it assumes that people eat the same food products, in the same quantities, and do not add any additional table salt, so consume less salt because of reformulation. It assumes industry is effective at reducing salt consumption across the range of targets.

The analysis took two separate steps, a static microsimulation of potential impact on salt reduction and a population modelling simulation for the health and healthcare cost impacts of these reductions in salt consumption.

### Sensitivity analyses

Three sensitivity analyses were included using point estimates and 500 iterations of Monte Carlo analysis.

1. A reduction in the effect to represent unintended public and industry response such as failure to achieve reformulation or the public adding more table salt was modelled, at 20% and 40% less than the main analysis across included products.
2. Alternate discount rates of 1.5% and 0% were applied to modelled outcomes as point estimates (consistent with the NICE’s recommended discounting comparators).

## Results

### Sample

A total of 586 adult participants aged 18 and over in the National Diet and Nutrition Survey were included. 2549 unique foods were reported to be consumed by this sample, of which 1532 were matched to a salt reduction target category (see Supplementary Table 1).

### Allocating products to salt reduction categories

It was possible to match 75 out of the 84 (89%) grocery food targets and 8 out of 24 (33%) of the out of home sector targets to their related foods.

### Salt consumption

If the government’s category-specific salt reduction targets set for 2024 were achieved in full, it is estimated that the adult population would see their salt intake reduce from 6.06g/day (95% CI: 5.18 – 6.31) to 4.94g/day (95% CI 4.73 – 5.15g), a reduction of 1.12g over 4 years (95% CI 1.05 – 1.20g) equivalent to 17.5% (**Table 1**). The reduction would be greater for males (1.34g (95% CI 1.22 - 1.47g)) than for females (0.93g (0.85-1.02g)). The distribution of salt consumption in the adult population at both baseline and scenario is shown in **Figure 2**.

**Figure 2:**
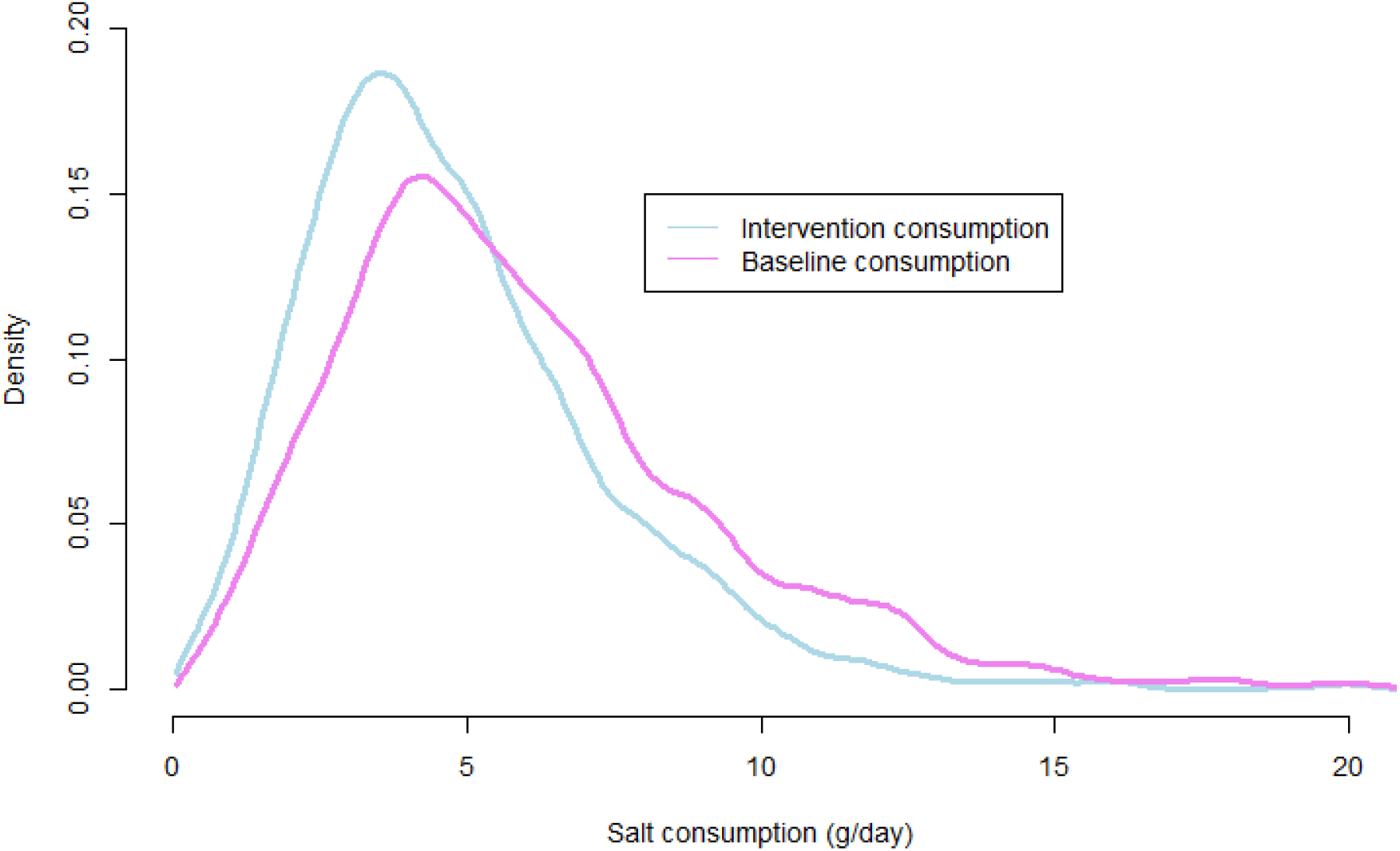
Survey-weighted distribution of salt intake at baseline and scenario.

### Disease burden and healthcare cost savings

Reduction in systolic blood pressure was calculated as 0.919mmHg (0.392 – 1.46mmHg) for females and 1.32mmHg (0.561 – 2.11mmHg) for males. In the first 20 years, this could avert 103,000 (95% UI 41,000 to 161,000) cases if of ischemic heart disease (IHD) and 25,000 (95% UI 10,000 to 39,000) of ischemic stroke across the population (full breakdown in **Table 2** below). Over the lifetime of the population, accounting for the potential for poorer health in later life, it would save 243,000 IHD- and stroke-related QALYs overall - 77,000 for women (95% UI 30,000 to 121,000) and 166,000 for men (95% UI64,000 to 263,000). The healthcare cost savings to the NHS are estimated to be £1.0 billion (95% UI £351m to £1.73 billion) over the population’s lifetime.

**Table 2.** Estimated impact of 2024 salt reformulation targets being met on reduction in quality adjusted life years (QALYs), disease burden, and healthcare costs. Interval represents the 2.5^th^ to 97.5^th^ uncertainty interval UI) generated from the Monte Carlo analysis.

| Outcome | Females | Males | Total |
| --- | --- | --- | --- |
| IHD cases (95% UI) | 26,000 (10,000-41,000) | 77,000 (30,000-119,000) | 103,000 (41,000-161,000) |
| Ischaemic stroke cases (95% UI) | 11,000 (4,000-17,000) | 14,000 (6,000-22,000) | 25,000 (10,000-39,000) |
| QALYs (95% UI) | 77,000 (30,000-121,000) | 166,000 (64,000-263,000) | 243,000 (94,000-383,000) |
| Healthcare costs (£million) (95% UI) | 323 (111-568) | 683 (240 – 1,153) | 1,001 (351-1,732) |

### Sensitivity analyses

The results of the first sensitivity analysis show that even if manufacturers reduced salt intake but missed the category targets by 20% or 40%, the reduction in the population’s salt intake would be less, but there would still be significant savings to QALYs and healthcare costs for males and females (see **Table 3**). These show reductions in potential effects approximately proportional to any shortfall in the desired impact of the targets on salt reduction, and that future discounted benefits are considerable so need accounting for in long-term decision-making.

**Table 3.** Results of sensitivity analyses 1 and 2 for 20% and 40% loss of effect of the intervention and for variations on discount rate to 3.5%, 1.5% and 0% for health and cost outcomes.

|  |  | Sensitivity Analysis 1: Effect reduction |  | Sensitivity Analysis 2: Discount rates |  |
| --- | --- | --- | --- | --- | --- |
|  |  | 20% | 40% | 1.5% | 0% |
| Salt reduction (g/day, 95% CI) | Females | 0.75 (0.68-0.81) | 0.56 (0.51-0.61) | *(As primary analysis)* |  |
|  | Males | 1.07 (0.97-1.18) | 0.81 (0.73-0.88) | *(As primary analysis)* |  |
| QALYs saved over 100 years (point estimate and 95% UI, thousands) | Females | 62 (22.8 – 99.5) | 46 (19.4 – 73.3) | 155 (62.9 – 226) | 285 (110 – 450) |
|  | Males | 133 (47.4 – 215) | 101 (39.7 – 159) | 329 (135 – 478) | 591 (224 – 920) |
| Healthcare costs saved over 100 years (point estimate and 95% UI, £millions) | Females | 260 (85.6 – 467) | 196 (67.5 – 343) | 578 (£203 – 1,000) | 957 (314 – 1,780) |
|  | Males | 549 (188 – 938) | 414 (148 – 716) | 1,200 (424 – 2,040) | 1,941 (548 – 3,320) |

## Discussion

This study shows that if the UK Government’s salt reduction targets for 2024 were met in full, it could lead to a 17.5% reduction in the salt intake of the adult population. This would prevent an estimated 103,000 cases of ischaemic heart disease and 25,000 ischaemic strokes over 20 years, leading to a saving of 243,000 QALYs and saving the NHS £1.00 billion over the population’s lifetime. Should the programme not meet its targets, for example a 20% or 40% shortfall, health benefits may be lost approximately linearly with these shortfalls. This study demonstrates that if implemented as intended, the UK salt reduction programme could have significant benefits on the population’s health. These effects are likely to be underestimated here given that limitations in both the NDNS data and salt reduction target category definitions mean that only 84 of the 108 target categories could be matched to foods reported in NDNS.

### Limitations

The National Diet and Nutrition Survey (NDNS) provides evidence on the UK population’s diet, but has several limitations. First, the majority of participants are white, with ethnic minority households under-represented (22). Second, the reliance on self-reported data means that consumption is often under-reported. We adjusted on the basis that the under-reporting of salt intake was proportional to urinary sodium, but it’s important to highlight that the urinary sodium survey also suffers from a small sample size, selection bias and incomplete sample collection. Also, whilst this adjustment will account for under-reporting of processed and pre-prepared foods, it will also account for salt added at the table and during cooking which is not covered by the targets. NDNS identifies foods that are purchased out of the home and they are coded as ‘takeaway’. However, there is significant underreporting of foods purchased as ‘takeaway’, with only eight of the 24 targets for the out of home sector matched to NDNS. The salt content (g/100g) of foods for NDNS Year 11 were taken from the Nutrient Databank (NDB) (23,24). While the sodium content of many key food categories, including bread, soup, crisps and breakfast cereals were updated in 2019, the sodium content of other categories may be out of date or rely on generic food composition data taken from the Composition of Foods Database (24). Any brand-specific reformulation that has taken place since will therefore not be captured in this study, meaning the health impacts reported may be over- estimated.

That some food categories could not be matched to their target products conferred a conservative bias on the estimate of salt reduction under these targets. It is not possible to meaningfully estimate to what extent this bias affects the estimates as the ambition of the targets and the quantities consumed of each product both vary greatly.

The scenario modelling relies on observational studies to link BP to disease outcomes (19) so unobserved confounding of effect sizes is possible. The PRIMEtime modelling structure assumes the risks of disease outcomes are independent, which may confer a conservative bias on results due to not accounting for superadded disutility and treatment costs associated with multimorbidity. In addition, only IHD and stroke are modelled here, while salt consumption is also potentially linked with diseases such as Chronic Kidney Disease, gastric cancer and dementia, conferring further conservative bias.

### Comparison to literature

This study is the first to estimate the potential public health impact of the Government’s current salt reduction policy and adds to the existing modelling evidence that population-level reductions in salt intake have the potential to lead to improvements in health outcomes. A previous study has estimated the cases of premature IHD and stroke averted based on 1) If the salt intake reduction of 1.0g/day between 2000 and 2018 was maintained to 2050 and 2) projected gains of reaching the WHO guideline of 5g/day per adult by 2030 (25). The study, using the same multi-state life table model structure (PRIMEtime) as in our study, found that 83,140 cases of IHD and 110,730 strokes could be prevented by 2050 if previous reductions in salt intake were maintained to 2050, equivalent to £1,260 million in healthcare savings (25). The salt reduction modelled in our study were slightly greater, particularly for males, and while the overall effects are comparable for IHD cases, they are somewhat lower for stroke cases.

The average reduction of systolic BP was modest, at 0.919mmHg and 1.32mmHg for females and males, respectively (26). This compares with an average treatment effect of 5.1mmHg for a first BP medication, though here the intervention effects many more people, over the entire life-course, avoids up-front costs to the NHS and is likely to reduce health inequalities.

Previous PRIMEtime modelling explored the potential impacts of the government sugar reduction strategy on health, with analogous voluntary reformulation targets. At 1.5% discount rates, this found the strategy could save 839,000 QALYs and £2.71bn over the lifetime (27). This study estimates potential benefits of less than half that for the sugar reduction strategy, though that paper highlighted that not all these benefits were likely to be seen as the government was aiming to achieve sugar reduction through weak assumptions, such consumers choosing healthier choices. Here, the opposite issue may be present – the inability to allocate all relevant products to their categories means some potential benefits of the programme may not be captured here.

### Policy implications

These results demonstrate the public health benefits that might be achieved if the salt reduction programme were successful. This study shows that the mean pace of salt reduction attributable to the salt reduction targets could be 0.28g/day per year over 4 years, and compares favourably to the previous success seen between 2003 and 2007 when the salt reduction programme was originally introduced, which achieved a rate of reduction of 0.175g/day per year (28).

This analysis was limited by related issues in that it was not possible to allocate many products to target categories, indicating potential weakness in monitoring progress towards the targets. Some target categories are overly specific, for example, the target for blue cheese (grocery category 4.4) is only relevant to products manufactured in the UK, which means many products are exempt from the target, but also that origin data is also needed for monitoring. The targets for battered or breaded chicken purchased out of the home are split based on energy content; under 200 kcal per serving, 200-400kcal and over 400kcal, making the data required to monitor them even more complex than the per 100g targets set for grocery foods.

If policymakers elect to stick with the voluntary programme, then more meaningful progress might be achieved by investing in better monitoring and more regular reporting. For example, including progress towards the salt targets as a metric that businesses should report on as part of the Food Data Transparency Partnership (FDTP) might lead to greater public and political scrutiny, and therefore more meaningful changes by manufacturers.

An alternative option is to make the targets mandatory, although enforcement of a mandatory scheme – for example, implementing a system to monitor and remove products from the shelves or issue fines for non-compliance - is likely to come at a high economic cost. Of the 62 countries that have salt reduction strategies worldwide, mandatory targets are in place in nine, with a further six having a combination of mandatory and voluntary schemes (29). Only three of these - Argentina, Paraguay and South Africa – have evaluated their mandatory schemes using labelling surveys led by independent academic groups(29). South Africa for example has implemented a two-phase mandatory salt target programme for 13 food categories that contribute the most to sodium intake, with interim targets introduced in June 2016 followed by stricter targets in June 2019 (30).

A 2021 survey found that 75% of products overall had a sodium content at or below the targets according to their labels, although there was great heterogeneity between categories (31). The same survey also found that for certain categories including bread and processed meat, the sodium content of products estimated from laboratory analysis was significantly higher than the values reported on the labels, which could overestimate compliance rates (31). Given the variation in salt strategies implemented and evaluation methods used across countries, it is hard to conclude whether mandatory targets are more or less successful than voluntary targets in achieving both compliance and population-level health benefits (31).

An alternative would be to introduce a tax on the salt content of foods to encourage reformulation, as recommended by the National Food Strategy (32) and the House of Lords in their report “Recipe for health: a plan to fix our broken food system” (33). This could be implemented as a scheme similar to the soft drink industry levy (SDIL), whereby food manufacturers are taxed based on the salt content of their products, with higher rates of tax imposed for foods with a higher salt content. The SDIL has been shown to lead to significant reductions in the sugar content of soft drinks, with the proportion of tax-eligible drinks (those containing 5g or more of sugar per 100ml) falling from 49% to 15% between September 2015 and February 2019 (34). It also generated over £1.0 billion in tax revenue between 2021 and 2024 (35). A flat rate tax might also be considered, with the National Food Strategy recommending a tax of £6 per kilo of salt sold as an ingredient for use in processed food or in catering and restaurants (32). While a salt tax may lead to a greater level of salt reduction than we’ve seen previously with voluntary reformulation, the food industry argue that it is harder to reduce salt levels in foods due to its technical properties, particularly as a preservative (36). Further research that assesses the benefits and limitations of different options, including the benefits and costs of different implementation mechanisms, needs to be addressed.

## Conclusion

This study found that should the 2024 salt reduction targets be achieved in full by the food industry, a major population-level reduction in sodium consumption and its associated disease burden would be predicted, with an estimated average of 17.5% less salt being consumed per person per day. This could be associated with 103,000 cases of IHD and 25,000 strokes in 20 years, accumulating to 243,000 QALYs and £1.0bn in healthcare cost savings over the lifetime of today’s adult population. Given these benefits, this study highlights the importance of prioritising salt reduction as part of a new focus on the prevention of ill-health. Policymakers should consider better monitoring and transparency of voluntary targets, making the targets mandatory or alternative approaches such as taxation which may lead to a faster rate of salt reformulation.

## Data Availability

Modelled outcomes data are available from the corresponding author. National Diet and Nutrition Survey data are freely available through the UK Data Service: https://beta.ukdataservice.ac.uk/datacatalogue/series/series?id=2000033

